# Seasonal forcing and waning immunity drive the sub-annual periodicity of the COVID-19 epidemic

**DOI:** 10.1101/2025.03.05.25323464

**Authors:** Ilan N. Rubin, Mary Bushman, Marc Lipsitch, William P. Hanage

## Abstract

Seasonal trends in infectious diseases are shaped by climatic and social factors, with many respiratory viruses peaking in winter. However, the seasonality of COVID-19 remains in dispute, with significant waves of cases across the United States occurring in both winter and summer. Using wavelet analysis of COVID-19 cases, we find that the periodicity of epidemic COVID-19 varies markedly across the U.S. and correlates with winter temperatures, indicating seasonal forcing. However, seasonal forcing alone cannot explain the pattern of multiple waves per year that has been so disruptive and unique to COVID-19. Using a modified SIRS model that allows specification of the tempo of waning immunity, we show that specific forms of non-durable immunity can sufficiently explain the sub-annual waves characteristic of the COVID-19 epidemic.

## Introduction

Much of the scientific and public discourse surrounding the COVID-19 epidemic and its waves has focused on the named “variants of concern” (VoC) (*1, 2*) that were dominate during each wave. While VoCs have played an unquestionably large role in the magnitude of the pandemic (*3*), the question of the timing of each wave is likely separate as we do not expect VoCs to emerge with a regular periodicity. The evolution of the SARS-CoV-2 virus may therefore have obscured the degree to which COVID-19 epidemic shows evidence of seasonality, in common with other respiratory viruses.

Many infectious diseases show pronounced seasonal patterns, producing waves of cases at particular times each year. Such seasonality is important for practical purposes in public health, allowing improved forecasting of heightened healthcare demand. Agents that transmit via the respiratory route are typically associated with a peak in the winter months, for reasons that may be due to climatic conditions and/or changing contact patterns during that period (*4*). In the case of the COVID-19 epidemic, waves of transmission have occurred not only in the winter, but also during the spring or summer, and have thus been out of sync with the classical pattern of respiratory viral pathogens. Untangling the seasonality of the COVID-19 epidemic from patterns in viral evolution, population immunity, the timing of government mitigation efforts, and epidemic stochasticity is a difficult yet imperative, task.

In the United States, multiple waves of infection have been documented each year since the start of 2020. Epidemic severity has tended to alternate between the northern and southern United States with evidence for a strong summer wave in the southern United States (*5*) as well as a negative temporal correlation between the number of cases and absolute humidity (*6*). This periodic regularity may be a separate phenomenon from the appearance of VoCs and so the precise patterns of seasonality, and thus their predictability going forward, remain to be determined.

While changes in case ascertainment make it hard to determine absolute incidence in the absence of large-scale community surveys (such as (*7*)), wavelet analysis offers an alternative approach to assess changing periodicity. Wavelet analysis is a spectral decomposition method similar to continuous Fourier transformation that is also able to handle non-stationary time series data and thus relevant in the pandemic context. Thus approach does not assume a single fixed periodicity and can therefore determine whether the oscillatory characteristic of the epidemic changes over time (*8*). For example, Grenfell et al. showed how the periodicity of measles cases in London changed after the onset of vaccination (*9*). Similar analyses of the COVID-19 epidemic for various countries around the world (*10, 11*), have as yet analyzed time periods in which only one or two epidemic waves had taken place and therefore offer limited insight into the long-term trajectory of the disease and the relative impact of VoCs.

The United States offers a useful case study for how climate effects epidemic seasonality, having a range of very different climates together with readily available estimates of case counts. Here we use wavelet analysis to determine the periodicity of COVID-19 waves in the United States using county level COVID-19 case estimates and determine how periodicity is correlated with measures of mobility, climatic, and demographic variables.

### COVID-19 displays both annual and sub-annual periodic signals

We applied wavelet decomposition to the logarithm of the 7-day rolling averages of cases per 100,000 individuals as compiled by the New York Times from January 21, 2020 through March 24, 2023, the entirety of the time they were reported (*12*). We analyze the periodicity of COVID-19 cases over that time-period using wavelet analysis – a technique to resolve a continuous time series into time-varying periodic components.

Wavelets of new infections at the state level were generally very stationary (for an example cases time-series and wavelet, see Fig. 1 showing the log cases, wavelet periodogram, and global wavelet spectrum for the state of Massachusetts), with the periodic signal often being comprised of two ridges – one roughly annual and one sub-annual (with a period in the range of 3-6 months). Within each state, this dynamic remained fairly constant across the entire testing period. Perhaps surprisingly, large-scale evolutionary events such as the first Omicron wave in late 2021 had little effect on each wavelet and the underlying periodicity of the infection dynamics. Instead, the evolutionary dynamics of SARS-CoV-2 and the immune evasion or increased transmission rate of the different VoCs seem to significantly affect the magnitude of the waves but otherwise fall within the periodic dynamics of the region.

**Figure 1:**
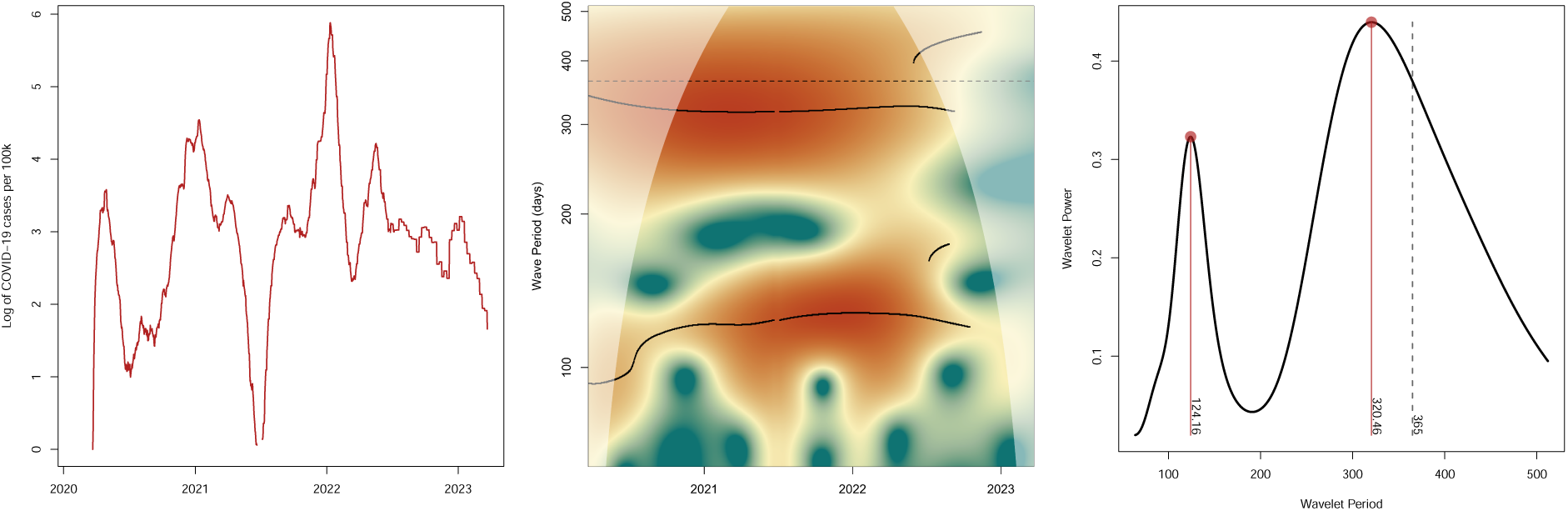
Massachusetts COVID-19 cases and periodicity. (**A**) The log COVID-19 cases per 100,000 residents in Massachusetts as estimate by the New York Times (*12*). (**B**) A wavelet periodogram of the log COVID-19 cases per 100k in Massachusetts. Color represents the power of the wavelet. The area with lighter coloring is outside the cone of influence and may be affected by edge effects. Significant ridges in the spectrum are shown as black lines. The two horizontal black lines represent stable components with periods of slightly less than a year and approximately four months. (**C**) The global wavelet spectrum for Massachusetts log cases per 100k, representing the average periodic components over the time-series.

As the wavelets are largely stable over this time period, we continue to analyze the global wavelet spectrum, which is calculated by averaging the wavelets over the entire time period and is analogous to a Fourier spectrum. The global wavelet is a distribution of the average periodic components that make up the time-series and is a simple summary of the periodicity of COVID-19 cases over the entire time period in question.

The state-level global wavelet spectra are relatively consistent across U.S. states and territories and generally show two major components [Fig. 2A]. The first has a period of approximately 365 days, as one would expect for a disease with classical winter waves of infections. The second major component is sub-annual and generally peaks between 120 and 180 days depending on the state and is consistent with the pattern of multiple large waves of transmission that occurred each year from 2020-2023 for most of the United States.

**Figure 2:**
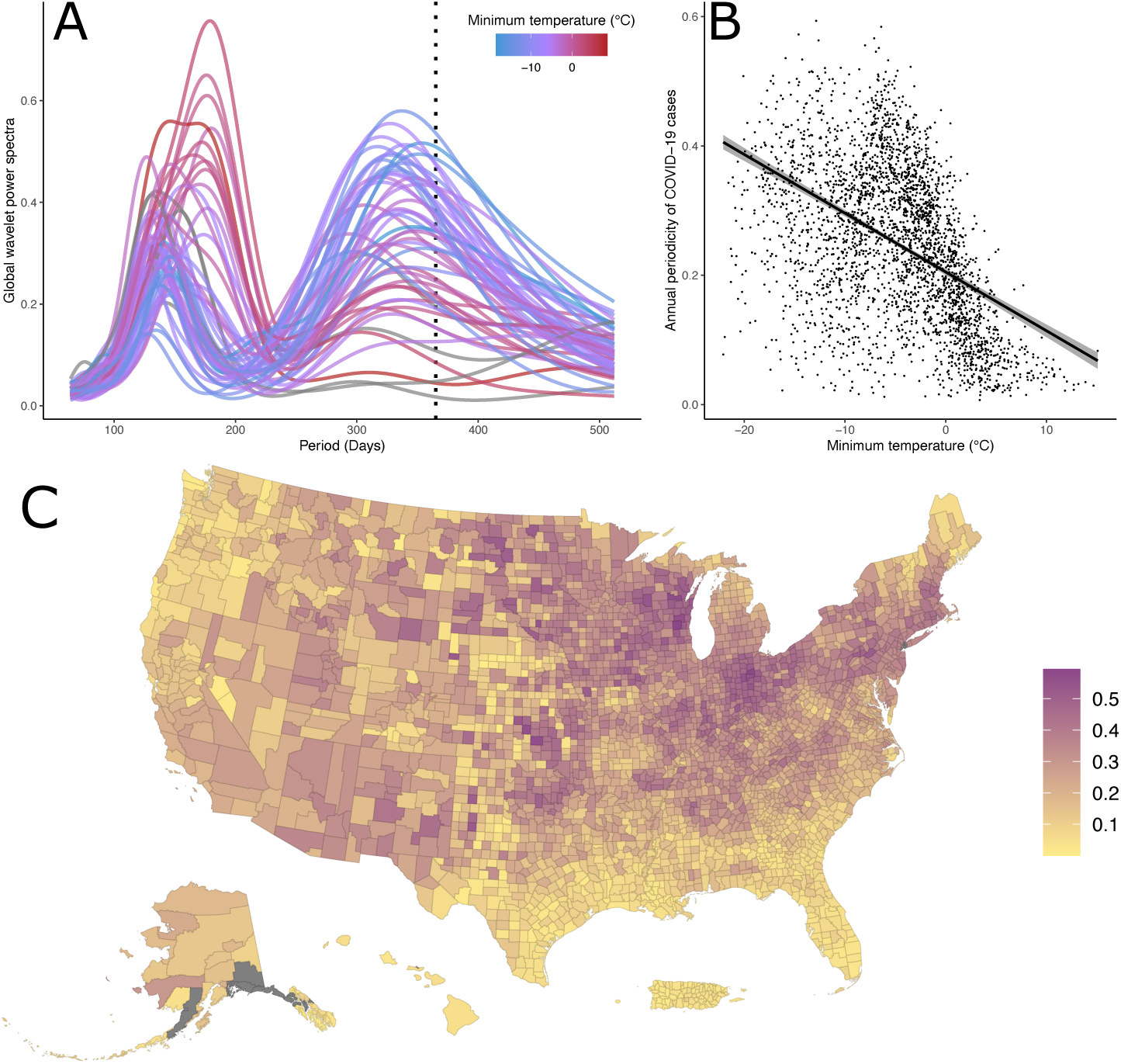
COVID-19 periodicity across the United States. Wavelet transformations were calculated for log of COVID-19 incidence for each U.S. state, Puerto Rico and the Virgin Islands. (**A**) Curves are colored by the minimum of the state-wide average monthly low temperature measured between 2020 and 2023. States with a warmer winter temperature are shown in red and cooler states in blue. Areas not included in NOAA’s historical climate dataset (Alaska, Hawaii, Puerto Rico, and the Virgin Islands) are shown in grey. A dashed line is shown at 365 days. (**B**) COVID-19 annual component as a function of temperature seasonality for each county in the United States. The magnitude of the annual component was calculated as the power of the global wavelet spectrum at 365 days. A linear regression and its 95% confidence interval are also shown. (**C**) A map of all counties in the United States colored by the magnitude of their annual component.

### COVID-19 periodicity is correlated to temperature

States with warmer winters (as quantified here by the average over 2020 to 2023 of the lowest of the monthly minimum temperature each year according to NOAA’s Monthly U.S. Climate Divisional Database (*13*)) were dominated by the sub-annual component and display little, if any, annual seasonality [Fig. 2A]. Conversely, states with colder winter low temperatures have annual and subannual components of similar magnitude in their global wavelet spectra and display a comparatively larger annual signal than states with warmer winters. Further supporting this pattern, of the four states and territories (Alaska, Hawaii, Puerto Rico, and the Virgin Islands – shown in grey) that are not included in the NOAA Climate Divisional Database, three have tropical climates and show essentially no signature of annual periodicity, while Alaska has a larger annual component and very cold winters. This difference in epidemic periodicity can be seen in the case counts as Florida, Louisiana, and Georgia (the states with the warmest winter climates) experienced two relatively similarly sized waves a year, while Minnesota, North Dakota, and South Dakota (the states with the coldest climates) experienced a large winter wave each of the three years with no or much smaller off-peak waves [fig. S4].

We extend this analysis to the county level and quantify the strength of the annual component as the power of the global wavelet spectrum at a period of 365 days. While this quantification is a relatively simple description of the seasonality of the epidemic in each county, it provides an intuitive way to summarize, in a single number, how strong the annual-scale periodicity of COVID-19 across the United States. In doing so, a clear pattern emerges. Counties in the southern and western United States, and particularly the southeast, show little annual periodicity, while counties in the Midwest and northeast of the United States experienced much more annual epidemics [Fig. 2C]. Counties around the Rocky Mountains have a less distinct pattern yet are also often less populous and therefore likely more dominated by stochastic dynamics.

In concert with this visually apparent trend [Fig. 2A], the power of the annual component for the COVID-19 epidemic across U.S. counties is significantly negatively correlated to temperature minimums (as defined by the average over 2020-2023 of the lowest monthly minimum temperature each year in the NOAA’s Climate Divisional Database (*13*)) across U.S. counties [Table 1, Fig. 2B]. Counties with colder winters were more likely to have experienced strongly annual COVID-19 epidemics, while counties with warmer winter temperatures were more likely to have epidemics dominated by sub-annual waves.

**Table 1:** Correlations between COVID-19 annual component and various climatic and socio-demographic variables. Wavelet analyses were calculated for the log of COVID-19 cases until March 2023 as estimated by the New York Times for each county (or county equivalent) in the United States. Regressions were calculated for the global wavelet transformation (average of the wavelet transformation over time) for a period of 365 days as a function of various climatic and socio-demographic variables. Positive slopes indicate that an increase in the variable in questions is correlated to an increase in the power of the annual component of COVID-19 in that county. Due to the large sample sizes, all correlations were statistically significant. Variables with *r*^2^
> 0.1 are bolded.

| Variable | Pearson's correlation | $r^2$ | source |
| --- | --- | --- | --- |
| **Minimum temperature** | -0.432 | **0.228** | (13) |
| **Maximum temperature** | -0.326 | **0.114** | (13) |
| **Difference of max and min temperature** | 0.354 | **0.166** | (13) |
| Average monthly precipitation | -0.113 | $2.89 \cdot 10^{-2}$ | (13) |
| Log population size | 0.207 | $2.09 \cdot 10^{-2}$ | (42) |
| % Population age 65+ | -0.130 | $9.02 \cdot 10^{-3}$ | (42) |
| % Population under 200% of the poverty line | -0.291 | $9.05 \cdot 10^{-2}$ | (42) |
| **% Population Insured** | 0.391 | **0.157** | (42) |
| Log population density | 0.157 | $8.86 \cdot 10^{-3}$ | (42, 43) |
| Log geographic area | $2.29 \cdot 10^{-2}$ | $1.63 \cdot 10^{-3}$ | (43) |
| Republican vote %, 2020 presidential election | $3.56 \cdot 10^{-2}$ | $5.75 \cdot 10^{-3}$ | (44) |
| Google mobility periodicity (retail) | 0.173 | $2.99 \cdot 10^{-2}$ | (45) |
| Google mobility periodicity (workplace) | -0.117 | $1.37 \cdot 10^{-2}$ | (45) |
| Log total number of COVID-19 cases | 0.199 | $2.66 \cdot 10^{-2}$ | (12) |
| **HHS region (ANOVA)** |  | **0.241** | (45) |

The maximum temperature of the county is also negatively correlated (to a lesser degree) with the strength of the annual component, a further link between colder climates and strongly annual COVID-19 epidemic dynamics [Table 1]. However, temperature variability (as defined by the difference between summer high and winter low temperatures) is positively correlated, possibly indicating that an annual epidemic cycle is associated with the overall extremity of the regional climate as well, rather than simple cold winters. Previous work on early pandemic dynamics has indicated higher transmission rates during the winter through direct correlations of cold temperatures with case counts (*6, 14*) and that the SARS-CoV-2 virus degrades more slowly in low temperatures and extreme relative humidity (*15*), a possible mechanism driving this periodic variability. However, the small a negative correlation with high temperatures in the summer may indicate that an annual epidemic cycle is associated with the overall extremity of the regional climate, rather than simply the cold winters.

In contrast with temperature, most socio-demographic characteristics of the county had little to no association with annual periodicity. Population size, population density, elderly population, geographic area of the county, and the political leaning of the county show little correlation to the COVID-19 epidemic periodicity [Table 1, fig. S6]. Thus, while the political partisanship of a county is known to be significantly correlated with the magnitude of the epidemic in that county (*16*), the timing of their waves was seemingly uncorrelated. We also, conducted similar wavelet analyses of Google mobility data (specifically their “retail and recreation” and “workplace” variables), that is both visually and intuitively periodic on an annual cycle (people move around more in the summer than winter). However, the mobility of the population explained under 3% of the variation in epidemic periodicity, compared to the more than 20% that is explained by temperature seasonality. Other variables that show relatively high correlation with the annual periodicity of the epidemic may be explained by their association with climate. For instance, of the ten Health and Human Services regions, the region of states around the Great Lakes (Region 5) has a significantly higher level of annual periodicity [fig. S6], a pattern that is consistent with temperature minimum. Similarly, the correlation with the percentage of the population with health insurance can be largely explained by low rates of insurance in Texas and Florida [Fig. S5], two states with comparatively warmer climates. A multivariable regression shows that the annual component is associated with both the minimum and maximum temperatures in a location (along with many of the other variables), further emphasizing the role of temperature in predicting stronger annual cycles [table S1].

While this work cannot establish the precise mechanisms underlying the periodicity we observe, we note that the regular timing of the winter peak each year [fig. S3] is generally consistent with the winter holiday period even in places with little evidence of an annual component (e.g., Florida) [fig. S4] and perhaps consistent with the increased mixing and changing contact patterns over the holiday period. However, holiday travel is relatively consistent across the U.S., yet COVID-19 periodicity is not and is instead correlated to temperature, supporting the possible climatic mechanism to seasonal forcing. Further comparisons with other regions around the world with different holiday schedules and climate variability may provide further evidence for this proposition. Most importantly, the positive correlation between the extent of climatic variability and annual periodicity is a strong indication that the observed periodicity is not simply due to chance arrivals of new SARS-CoV-2 strains at approximately annual intervals, and instead suggests that some climate-related drivers (including behavioral consequences) are an important driver of annual seasonality. A prediction of the hypothesis that seasonal forcing drives the annual periodicity is that this periodicity should be stronger where seasonality is stronger. Our findings are consistent with this finding, and would be hard to explain parsimoniously if the annual period were a byproduct of chance timing of introduction of new variants.

### A model of sub-annual periodicity considering individual waning immunity

While the previously described wave decomposition analyses provided insight and intuition into the character of the periodicity of the COVID-19 epidemic, these analyses are non-mechanistic. Previous theory on seasonal forcing shows how epidemics can be forced into an annual cycle or longer multiennial cycles (*9, 17, 18*). For instance, Dushoff et al. show how undetectably small changes in transmission rate over the course of an annual cycle can result in large oscillations in influenza incidence through dynamical resonance of the disease and seasonal dynamics (*19*). However, this seasonal forcing theory largely pertains to epidemics with annual or longer cycles.

The classical SIRS model (Susceptible-Infectious-Recovered-Susceptible) can drive waves in the number of infected individuals through cycles of the depletion and replenishment of the number of susceptibles as immunity in the population wanes. However, the classical SIRS model does not consider partial or waning immunity, but absolute immunity that wanes immediately (with the time persons are immune distributed exponentially in the population) (*20*). Under these classical assumptions, while the inter-epidemic period can be controlled as a function of the average length of time an individual is immune, epidemic waves are always damped oscillations (decrease in size from one wave to the next) and are thus not maintained over long periods (*21*). The first years of the COVID-19 pandemic were characterized by a series of waves of new infections of equal or successively greater magnitude, a pattern that cannot be explained by the classical SIRS model.

To provide intuition into possible mechanisms that could be driving this more unusual sustained sub-annual periodic behavior, we introduce a generalization of the classic SIRS epidemic model that includes both partial immunity that wanes over time as well as seasonal forcing [Fig. 3]. Immunity to reinfection is known to wane (*22, 23*) and seasonal forcing is a significant periodic driver in other respiratory pathogens (*17, 24*). Here we prove whether one or both of these mechanism can explain the sub-annual periodicity we have described.

**Figure 3:**
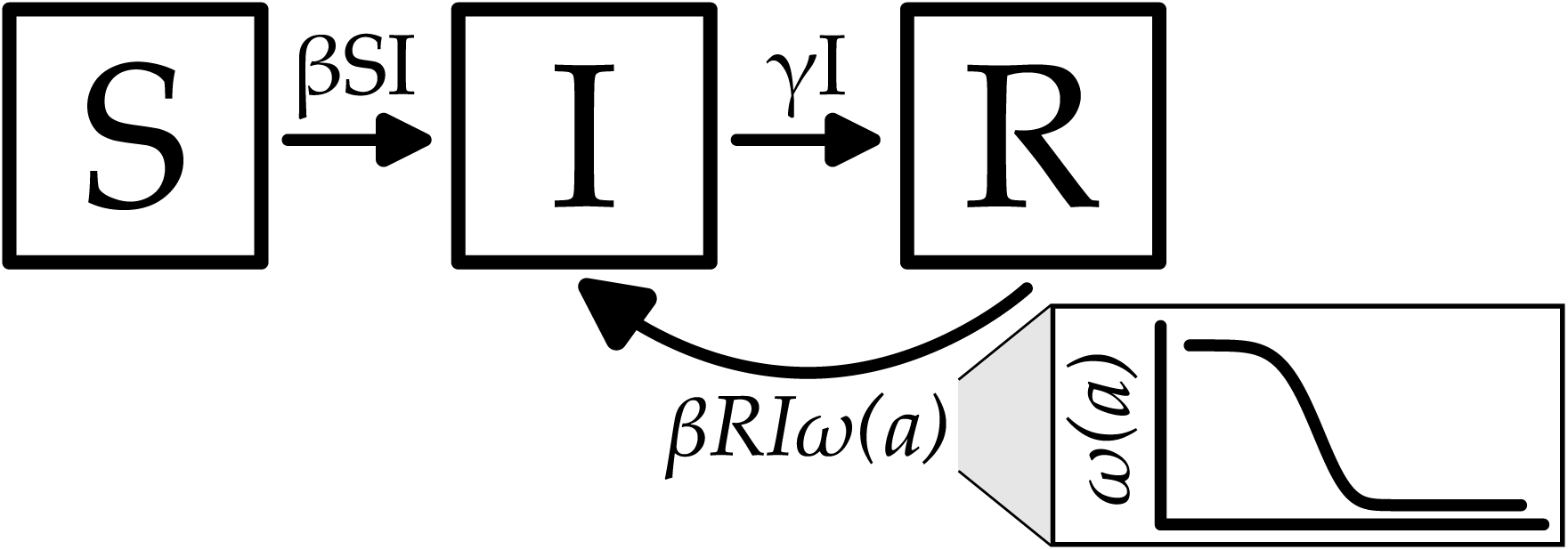
SIR model with waning immunity compartment diagram. A representation of the proposed SIR model with waning immunity. *S*, *I*, and *R* represent the susceptible, infected, and recovered states respectively and are all functions of time, *t*. *β* is the transmission rate and may be either a static variable or a function of time, *t*, while *γ* is the rate of recovery from illness. In the classical SIRS model, recovered individuals lose all immunity and return to the susceptible compartment after some time. In the model presented, individuals remain “recovered” for perpetuity but over time can be reinfected as a function of waning immunity *ω*(*a*), where *a* is the time since recovery. In this model *S* represents an initial state made up of individuals who have never been infected and have no immunity.

The model can be described as a system of partial differential equations with Susceptible (*S*), infectious (*I*), and Recovered (*R*) classes that are a function of time (*t*) and time since recovery from infectiousness (*a*). Individuals can become infected from both the Susceptible and Recovered classes with immunity from infection for the Recovered class described by a function *ω*(*a*). While this function can take any form, here we use a functional form based on the cumulative distribution function for a Weibull distribution that allows for easy tuning between exponentially and more stepwise (sigmoidal) decaying immune waning functions [e.g., see Fig. 3 and fig. S7]. The model is as follows:

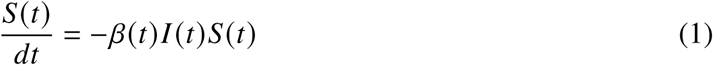

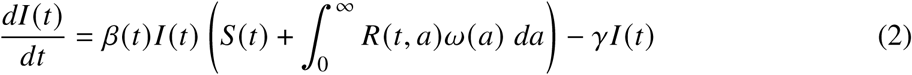

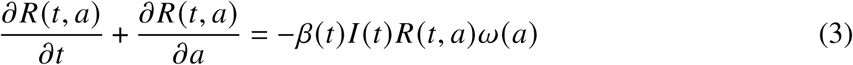

with the following boundary condition:

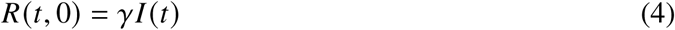

and the following waning immunity function:

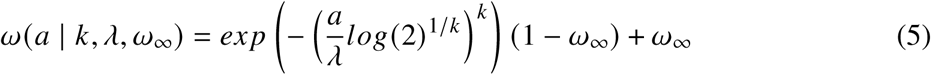

where *γ* represents the recovery rate from the infected to the recovered (immune) class and *β*(*t*) is the transmission rate at time *t* (*25*). The immunity waning function is governed by three parameters: *k* is the shape parameter that controls the steepness of the waning, *λ* represents the “half-life” and controls the timing, and *ω*_∞_ is the asymptote or amount of immunity maintained for perpetuity. For a more detailed discussion of the model including the sinusoidal seasonal forcing function, see materials and methods.

For computational simplicity, we simulated an individual-based version of the SIR model with waning immunity and seasonal forcing. We ran 125,000 simulations using Latin hypercube sampling (*26*) to sample the three immune waning parameters with different amounts of annual seasonal forcing as part of the transmission rate. The simulations were then analyzed based on the periodicity of newly infected individuals as defined by the average inter-epidemic period between waves in order to understand which immunity regimes generate sub-annual epidemic periodicity that is maintained over a relatively long period of time (i.e., effectively undamped oscillations) with and without seasonal forcing. Annual periodicity is defined as any simulation with an average inter-epidemic period of between 9 and 18 months and sub-annual of less than 9 months, both requiring more than 5 total waves during the 10 simulation years to filter out strongly damped oscillations. Simulations that did not result in either annual or sub-annual waves either reached a stable equilibrium endemic state or resulted in waves less frequent than every 18 months.

The model is able to produce sub-annual waves when three conditions are met: there is little long-term immunity (*ω*_∞_ < 0.5), immunity wanes relatively sharply (*k* > 1.5), and half-life time of immunity is short (*λ* < 200 days) [fig. S7]. In practical terms, this represents waning functions that have a sigmoidal shape and thus a period of strong immunity, a transition period with partial immunity that occurs with about half a year or less, and very little immunity in the long-term. This durability is roughly in line with epidemiological estimates of the durability of immunity to SARS-CoV-2 (*22, 23, 27*).

In general, the shape parameter of the waning function (*k*) and the amount of long-term immunity (*ω*_∞_) determine whether stable periodicity is possible, while the frequency of that periodicity is largely a function of the half-life of the immunity (*λ*). For example simulation trajectories and immune waning functions, see figure S7. Adding seasonal forcing to the model can create more variable and chaotic dynamics than the damped or stable waves generated from just waning immunity [for example trajectories with seasonal forcing see fig. S9] as the two periodic generating mechanisms (seasonal forcing and waning immunity) may resonate or interfere with each other.

Perhaps most importantly, including annual seasonal forcing on the transmission rate had little to no effect on the generation of sub-annual periodicity and instead only increased the likelihood of annual waves [fig. 4]. This is still true if only the first four years of the simulation are considered [fig. S8] to match the period of COVID-19 case counts we analyzed in the previous section. Thus, while seasonal forcing undoubtedly would impact the magnitude and timing of the epidemic waves, likely in a chaotic way due to the resonance and synchrony of the two processes (*28, 29*), it is unlikely to change the importance of non-durable immunity in generating the such rapidly reoccurring epidemic waves unusual to the COVID-19 epidemic.

**Figure 4:**
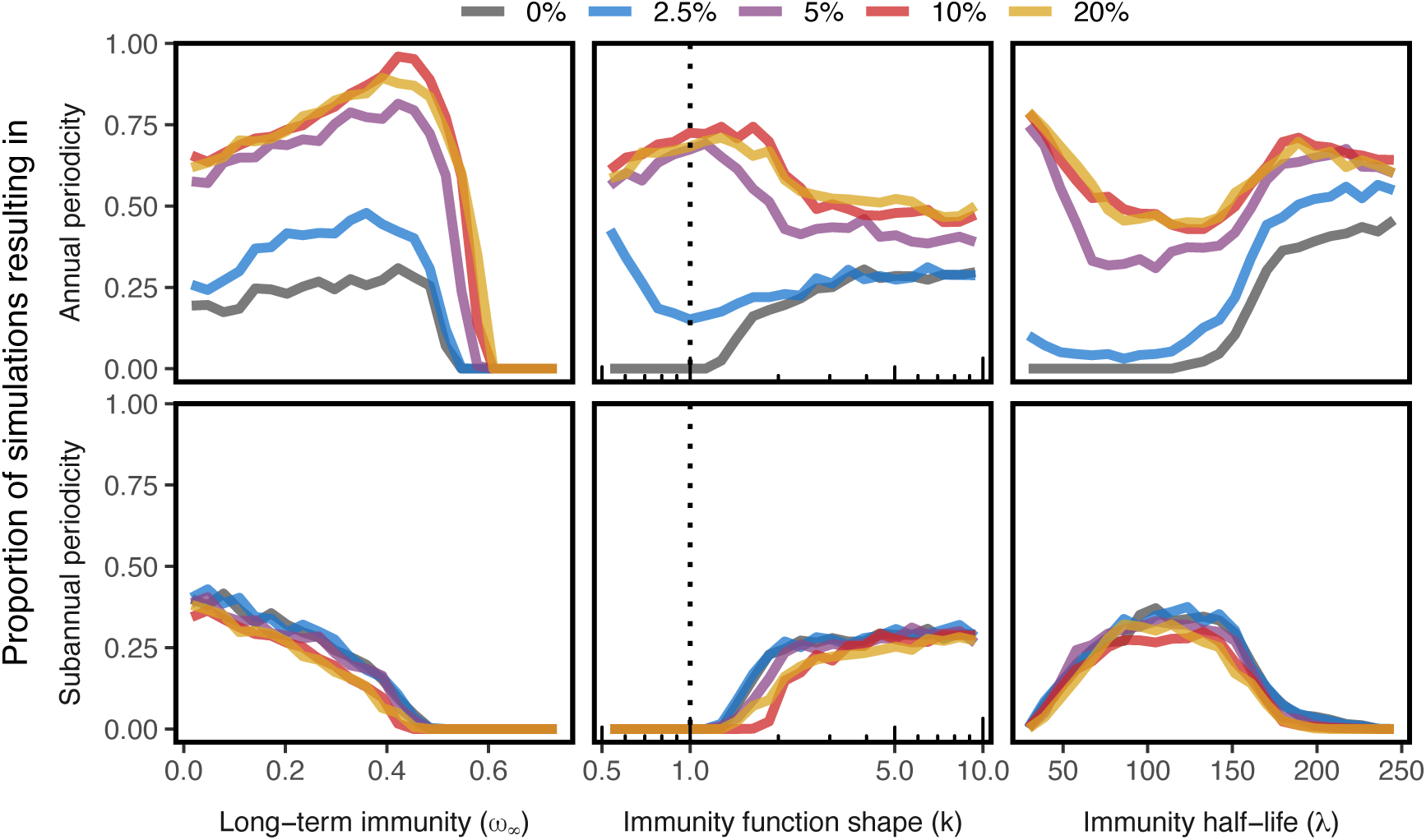
Seasonal forcing does not markedly affect sub-annual periodicity in simulated epidemics. We ran 125,000 simulations of the individual-based implementation of the SIRS model with individually waning immunity, with 25,000 simulations for each of five different parame-terizations of seasonal forcing (shown in color with *θ* = 2.5%, 5%, 10%, and 20%, or when the peak transmission rate is *θ*% higher transmission than at its lowest). Immune parameters were chosen randomly with Latin Hypercube Sampling for each seasonality regime (*ω*_∞_ ∈ (0, 0.75), *k* ∈ (0.5, 10), and distributed logarithmically *λ* ∈ (25, 250)). Annual periodicity is defined as any simulation with an inter-epidemic period of between 9 and 18 months and sub-annual of less than 9 months, both requiring more than 5 total waves during the 10 simulation years to filter out strongly damped oscillations. Simulations that did not result in either annual or sub-annual waves either reached a stable equilibrium endemic state or resulted in waves less frequent than every 18 months. The probability of an annually recurring epidemic generally increases with increasing seasonal forcing, dependent on the immunity characteristics. However, the presence of seasonal forcing has little to no effect on driving sub-annually reoccurring epidemics. The dashed lines represent exponentially waning immunity, a good proxy for the classic SIRS model, and where undamped sub-annual waves are never generated. For an epidemic to be periodic at all immunity must wane sharply (*k* ≳ 1.5) and the long-term immunity must be marginal (*ω*_∞_ ⪅ 0.5).

However, simulations with increased seasonal forcing do result in an increased probability of annual periodicity, with no accompanying reduction in the chance of sub-annual periodicity. This increase in annual periodicity possibly provides an explanation for the temperature-mediated periodic patterns we described earlier in which areas of the United States with colder winters – and thus likely stronger seasonal forcing – showed a dominate annual signal in COVID-19 cases.

The evolution of the SARS-CoV-2 virus has unarguably played a large role in the magnitude and duration of each wave of infections, yet evolution depends on the inherently stochastic nature of mutation while seasonality does not. Nor is it easy to explain how evolution alone can produce periodic behavior that is associated with climate (though see (*30, 31*) for a possible theoretic basis for evolution induced waves). The regular timing of the waves that we have described implies a more deterministic mechanism (such as immune dynamics or seasonal forcing) has been a necessary driver of the periodic dynamics. While previous work finds that short-duration infections, continuous antigenic drift, and complete cross-immunity between strains can generate periodic waves of infections (*30, 31*), here we show short-term immunity that wanes quickly is a sufficient condition for regular sub-annual periodicity, independent of multi-strain dynamics (see (*32*) for a more in-depth discussion of reinfection rates as function of recovery time).

Moreover, antigenic evolution effectively accelerates the rate with which the immunity of the population wanes as replacing strains out-compete their predecessors, and so the emergence of a VoC able to invade existing neutralizing antibodies has a similar effect to a sharply declining waning immunity function. Therefore, the effects of continual evolution and a waning immune response to a specific strain are difficult to practically separate and measure in natural populations. An individual’s “effective immunity” does not only consider their individual immune response, but also the whether the individual immune response is well matched to the virus that is circulating at that time. Thus, we are not suggesting individual immune response dynamics instead of evolution as the sole mechanism of periodic dynamics, but that the recruitment of susceptible individuals – whether through antigenic drift or the waning of an individual immune response (or some other demographic process) – is a necessary component of periodically reoccurring epidemics and that a sharply waning “effective” immunity is sufficient to explain the sub-annual periodicity we have previously described.

Previous work has examined periodic disease dynamics that are caused by or interacts with the repeated turnover or coexistence of viral strains (*33, 34*). Continued theoretical work aimed to understanding how large-scale evolutionary change in viral antigens – such as that which occurred in the BA.1, BA.2, and BA.2.86 variants (*35,36*) – interacts with the immunity-mediated sub-annual periodic dynamics described here is still warranted.

Just as evolution or waning immunity are unlikely to be correlated with climate, seasonality alone cannot explain the sub-annual periodicity we have shown, but it can emphasize an annual component [Fig. 4. The stronger signal for annual periodicity we see in regions of the United States with colder winters may therefore be the result of these factors operating together, with waning immunity driving the sub-annual dynamics and strong seasonal forcing masking it in favor of a more annual epidemic in places with cold winters.

## Conclusions

While the patterns reported in this work have been consistent throughout the study period, the accumulation and loss of immunity, unforeseeable and stochastic viral evolution, and unpredictable nature of complex and possibly chaotic wave dynamics make detailed forecasting a challenge. While some have postulated that COVID-19 will inevitably transition to annual winter waves (*24*) that are a classical characteristic of other respiratory viral pathogens in the endemic state (*17,19,24,37,38*), we have shown here that this transition has yet to take place, though annual waves have been a key part of the dynamics, especially in colder climates. Continued surveillance will be essential to monitor if these dynamics have stabilized into a predictable cycle or continue to evolve. As the immediate threat of the pandemic has dwindled, mild cases are likely to remain under-counted and present a challenge. While wastewater surveillance emerged during the pandemic as a new means to track disease trends in close to real time (*39, 40*), it has not been universally applied and has remaining theoretical challenges as a direct replacement for high-quality estimates of disease incidence. Despite these challenges, more current wastewater surveillance data suggests that the patterns of regular winter waves of new cases across the United States and less consistent “off cycle” wave may still be relevant (*41*), reinforcing the importance of understanding the periodicity of the disease as a tool for predicting dynamics going forward. Nevertheless, a thorough analysis of wastewater surveillance data collected from diverse regions going forward would allow us to understand if the patterns observed here persist in the coming years.

We have shown evidence for distinct patterns of seasonality in COVID-19 cases across the U.S.,correlated with climate. While evolution is known to have played a large role in the transmission and immune escape of the SARS-CoV-2 virus (*3*), here we show that the periodicity of the COVID-19 epidemic remained relatively stable during the pandemic period despite large evolutionary events and is correlated to temperature lows. In warmer regions circulation throughout the year is marked by multiple sub-annual waves occurring every 3-4 months, while in regions with colder winters, an annual component predominates. We also show that annual seasonal forcing alone is unable to explain sub-annual waves but can produce such dynamics in concert with another periodic generating mechanism such as the steeply waning, short-term convalescent immunity proposed here. Although we are not able to disentangle exactly how viral evolution combines with the waning human immune response, we note that antigenic evolution produces accelerated waning. Improved understanding of how seasonal forcing and viral evolution interact with quickly waning immunity will permit future extensions of the classical SIRS model to describe other periodic epidemics.

## Data Availability

All data used in this study is publicly available and the sources are cited in the manuscript. All code for both the spectral analysis and model simulations is
available at https://github.com/ilanrubin/COVID-19-Seasonality.

## Acknowledgments

We would like to thank colleagues from the Harvard T.H. Chan School of Public Health, Center for Communicable Disease Dynamics for their helpful and thoughtful discussion and feedback throughout the writing of this paper.

## Author contributions

I.N.R. conceived paper and prepared initial draft. I.N.R and W.H. prepared further drafts and all authors contributed to the final draft and provided approval. All authors contributed to model and analysis methodology. I.N.R. and M.B. performed programming and software development. I.N.R created visualizations and performed analyses.

## Data and materials availability

Code for both the spectral analysis and model simulations is available at https://github.com/ilanrubin/COVID-19-Seasonality.

