## Supplemental Materials for "Seasonal forcing and waning immunity drive the sub-annual periodicity of the COVID-19 epidemic"

### Materials and Methods

#### Periodicity Analysis of COVID-19 Incidence

We generated wavelet transformations of COVID-19 incidence using the package *WaveletComp* (*46*) in R (*47*). We analyzed state and county-level log transformed 7-day rolling averages of new cases per 100,000 individuals as compiled by the New York Times from January 21, 2020 through March 24, 2023 (*12*). Only counties with more than 500 total COVID-19 cases were included in the analysis to eliminate the highly stochastic nature of epidemics in small populations.

We then analyzed the resulting power spectra in the context of historical climate data (monthly average temperature highs and lows compiled by the National Oceanic and Atmospheric Administration's Monthly U.S. Climate Divisional Database – NClimDiv) (*13*), population demographic information base on the 2020 United States Census and American Community Survey (*42, 43*), Google's Community Mobility Reports (*45*), and results of the 2020 presidential election (*44*). For more detailed information on data sources and methodology, please see below.

Global wavelet spectra are defined as the average of the wavelet spectra over the entire course of the time series. The strength of the annual component of a global wavelet spectrum is used to quantify the magnitude of annual-scale periodicity in the epidemic and defined as the value of the global wavelet spectrum at exactly 365 days.

#### Model with Waning Immunity

While the previously described wave decomposition analyses examine the periodicity of the COVID-19 epidemic, these analyses are non-mechanistic. The classic model used to provide intuition into possible mechanisms that could be driving this sub-annual periodic behavior, is the SIRS (Susceptible-Infectious-Recovered-Susceptible) compartment model. In this model, the population is divided into and moves between the Susceptible ($S$), Infectious ($I$), and Recovered ($R$) classes. It is commonly written as the following set of three ordinary differential equations:

$$\frac{S(t)}{dt} = -\beta(t)I(t)S(t) + \omega R(t) \tag{S1}$$

$$\frac{dI(t)}{dt} = \beta(t)I(t)S(t) - \gamma I(t) \tag{S2}$$

$$\frac{dR(t)}{dt} = \gamma I(t) - \omega R(t) \tag{S3}$$

where $\beta$ is the transmission rate, $\gamma$ is the recovery rate, and $\omega$ is the rate of loss of immunity. Immunity in the SIRS model is completely sterilizing and the time individuals are immune is exponentially distributed with a mean of the inverse of the rate individuals revert to susceptible (i.e., the mean time immune = $\frac{1}{\omega}$). Periodicity in the SIRS model is driven by a return of individuals from the Recovered (aka immune) class to being susceptible.

Here we introduce a generalization of the classic SIRS epidemic model that includes individual partial immunity that wanes over time. This model can be described as a system of partial differential equations with the same Susceptible ($S$), infectious ($I$), and Recovered ($R$) classes, yet here they are a function of both time ($t$) and time since recovery ($a$). Individuals can become infected from both the Susceptible and Recovered classes with immunity from infection for the Recovered class described by a function $\omega(a)$. The proposed model is as follows:

$$\frac{S(t)}{dt} = -\beta(t)I(t)S(t) \tag{S4}$$

$$\frac{dI(t)}{dt} = \beta(t)I(t)\left(S(t) + \int_0^\infty R(t,a)\omega(a)\, da\right) - \gamma I(t) \tag{S5}$$

$$\frac{\partial R(t,a)}{\partial t} + \frac{\partial R(t,a)}{\partial a} = -\beta(t)I(t)R(t,a)\omega(a) \tag{S6}$$

with the following boundary condition:

$$R(t,0) = \gamma I(t) \tag{S7}$$

$\gamma$ represents the rate of recovery from the infected to the recovered, or immune, class $\beta(t)$ is the transmission rate at time $t$ . $\beta(t)$ can be a scalar such that the transmission rate is fixed and independent of time or season. Alternatively, annual seasonal forcing can be modeled as a sinusoidal transmission function $\beta(t) = \beta_0(1 + \theta \cos((2t/365 + s_\theta)\pi))$, where $\beta_0 \in \mathbb{R}^+$ is the baseline transmission rate, $\theta \in [0, 1]$, represents the amplitude of seasonal forcing and $s_\theta \in [0, 1)$ represents a phase shift parameter that governs the time during year that peak transmission occurs. Any other functional form can also be used for $\beta(t)$ in place of the sinusoidal forcing.

This model allows for explicit control of the shape of the individual immune waning function, $\omega(a \mid k, \lambda, \omega_\infty)$, that is a function of the time since recovery from a previous case of COVID-19 infection ($a$). The individual waning function can take any form. Here we propose and analyze the following function:

$$\omega(a \mid k, \lambda, \omega_\infty) = exp\left(-\left(\frac{a}{\lambda}log(2)^{1/k}\right)^k\right)(1 - \omega_\infty) + \omega_\infty \tag{S8}$$

The proposed function is loosely based on the Weibull cumulative distribution function and allows for a large variety of biologically relevant scenarios with few parameters. The waning immunity function wanes from 1, indicating full immunity, to $\omega_\infty \in [0, 1]$ that represents the long-term immunity (when $\omega_\infty = 0$ immunity is eventually completely lost). The rate of waning governed by a shape parameter $k \in \mathbb{R}^+$ and a timing parameter $\lambda \in \mathbb{R}^+$. Large $k$ represent a waning function that decreases from full immunity to $\omega_\infty$ in a stepwise manner, while smaller $k$ represents more gradual waning. The timing parameter $\lambda$ represents the "immunity half-life," or the time when immunity is halfway between 1 and $\omega_\infty$. If $k = 1$ and $\omega_\infty = 0$ the waning function describes exponential waning with an exponential decay rate of $1/\lambda$.

For examples of what this waning immunity function can look like with different parameterizations and the resulting dynamics, please see Figure S7.

### Individual-based simulations

We simulated an individual-based approximation of the proposed PDE model with discrete time (time resolution of a day). This was done for computational and programmatic ease. Individual immunity is tracked deterministically for each individual who has recovered from symptomatic disease. The number of susceptible individuals exposed to the disease each day is calculated as a binomial distribution with probability $p = 1 - exp(-I * R_0 * \gamma * \beta(t)/N)$ The code is available in an online repository.

Parameterization of the fixed parameters was chosen to roughly mimic that of COVID-19. As a relatively simple compartment-based model, it is not intended to precisely fit actual epidemic curves, but rather as a toolbox able to reproduce the general dynamics (e.g., sustained sub-annual periodicity) with a few, easily tunable parameters. Thus, the specific parameterization is intended to be feasible but not based on specific data. The most important parameters used in simulations are show in Table S2.

The individual based implementation is stochastic in nature, but when run on a population size of 1,000,000, resulted in sufficiently deterministic-like dynamics. Each simulation with the chosen parameterization was run for 3650 days. In order to eliminate the effect of the stochastic extinction of the disease, the simulation is reseeded with a single newly infected individual anytime it goes extinct.

After the simulation was completed, we counted the peaks in number of infected individuals. Peaks were identified with a recursive algorithm and defined as any point that is a local maximum and has a 1% prominence (the number of infected individuals are at least 1% higher at the local maxima than the minimum point between it and the next closest peak) or the simulation start or finish if there is no peak between it and one end of the time series. This is a pragmatic definition of an epidemic peak, but one we feel has more relevance to real-world epidemic scenarios than analytical determinations based on the differential equations as it dismisses any deterministically generated epidemic peaks that are too small to have practical significance as well as local maxima caused by stochasticity.

Annual periodicity is defined as any simulation with an average inter-epidemic period of between 9 and 18 months and sub-annual of less than 9 months, both requiring more than 5 total waves during the 10 simulation years to filter out strongly damped oscillations. Simulations that did not result in either annual or sub-annual waves either reached a stable equilibrium endemic state or resulted in waves less frequent than every 18 months.

### COVID-19 case data

COVID-19 case counts were taken from the New York Times' COVID-19 case estimates (*12*). Cases were converted to cases per 100,000 individuals to control for population size between municipalities. The cases per 100k were then log transformed before spectral decomposition analysis to limit the out-sized effect of the initial Omicron wave in the winter of 2021-2022 and to emphasize the timing, rather than the magnitude of the waves.

### Climate data

Historical temperature and precipitation data for each county were obtained from the National Atmospheric and Oceanic

### Population demographic data

We obtain estimates of the population size, elderly population, population in poverty, and population insured for each county from the 2020 U.S. Census American Community Survey (*42, 43*).

### Mobility data

To estimate changes in mobility we use Google's mobility dataset (*45*). We specifically use the "workplaces" and "retail and recreation" mobility variables as the two variables with the least sparse reporting. The mobility variables are presented as a percentage change compared to the baseline and "show how visits and length of stay and different places change compared to the baseline" (*45*). The baseline is calculated during the 5-week period from January 3, 2020 through February 6, 2020.

To analyze the periodicity of the mobility variables at each location we perform the same wavelet decomposition analysis that we do on the COVID-19 case data. Similarly to with COVID-19 cases, the annual periodicity of a county's mobility is then defined as the magnitude of the global wavelet spectrum at exactly 365 days. To ensure a reasonable estimate of the periodic signal, only counties with at least 500 days of mobility data were included in this analysis.

Notably, mobility is not equivalent to interaction between individuals, so likely does not capture the extent of the changing social network structure and increased contacts during the holiday season each year.

### Election data

We define the political leaning of each county as the percentage of voters in each county that voted for a republican candidate during the 2020 presidential election. Election data is sourced from The New York Time's 2020 election dataset (*12*).

### Mapping data

The shapefiles used to map all county-level variables were obtained from the U.S. Census Bureau's 2020 Gazetteer Files (*43*).

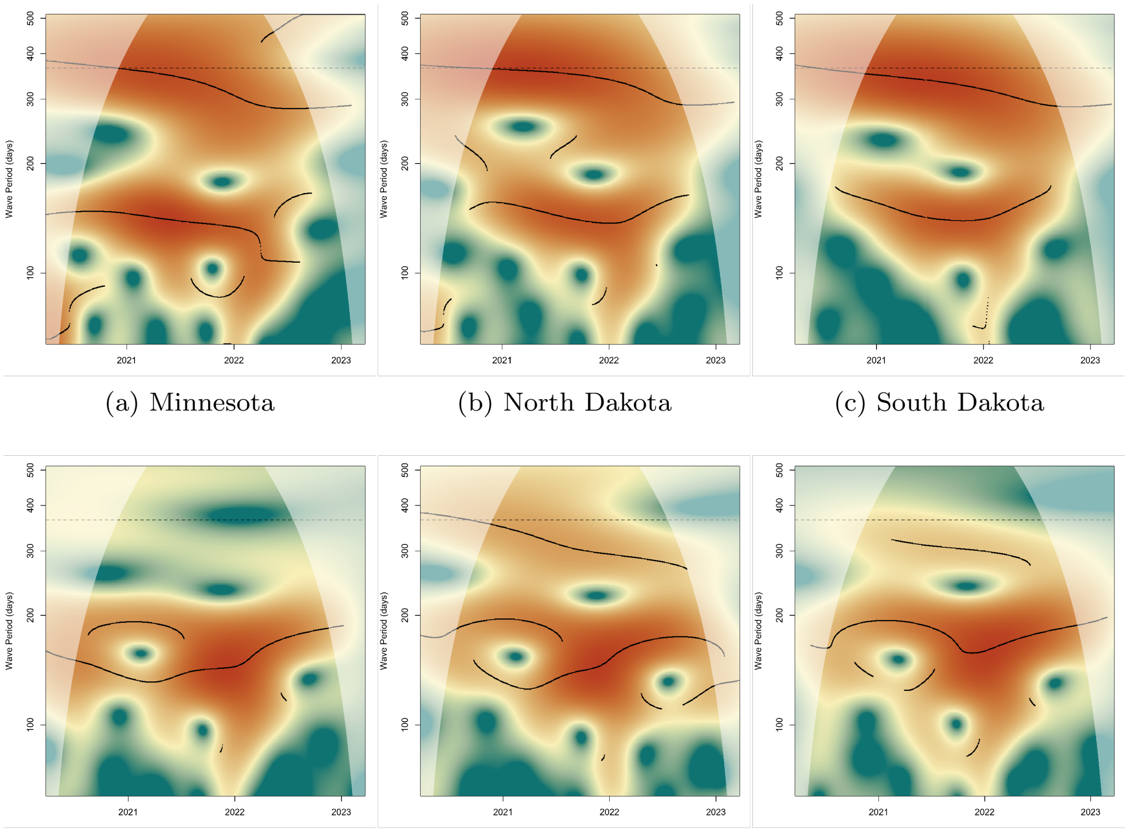


(a) Minnesota (b) North Dakota (c) South Dakota

(d) Florida (e) Georgia (f) Louisiana

**Figure S1**: **Wavelets of COVID-19 for the states with the coldest (a-d) and warmest (d-f) winters climates.** Wavelets the log of the number of cases of COVID-19 per 100k are shown for the three states with the coldest winter climates (Minnesota, North Dakota, and South Dakota) and the warmest winter climates (Florida, Georgia, and Louisiana). Black lines depict ridges in the wavelet. Wavelets were relatively stable throughout the period of study allowing for us to simplify the analysis to use the global wavelet transformations. Global wavelet transformations for all 50 states are shown in fig. S2. Historical temperature data comes from NOAA's Monthly U.S. Climate Divisional Database. Colder regions tended to have larger winter waves of new infections and little if any disease in the summer. Conversely, the warmer regions had large summer waves of new infections each year that were of similar or even larger magnitude to their winter waves.

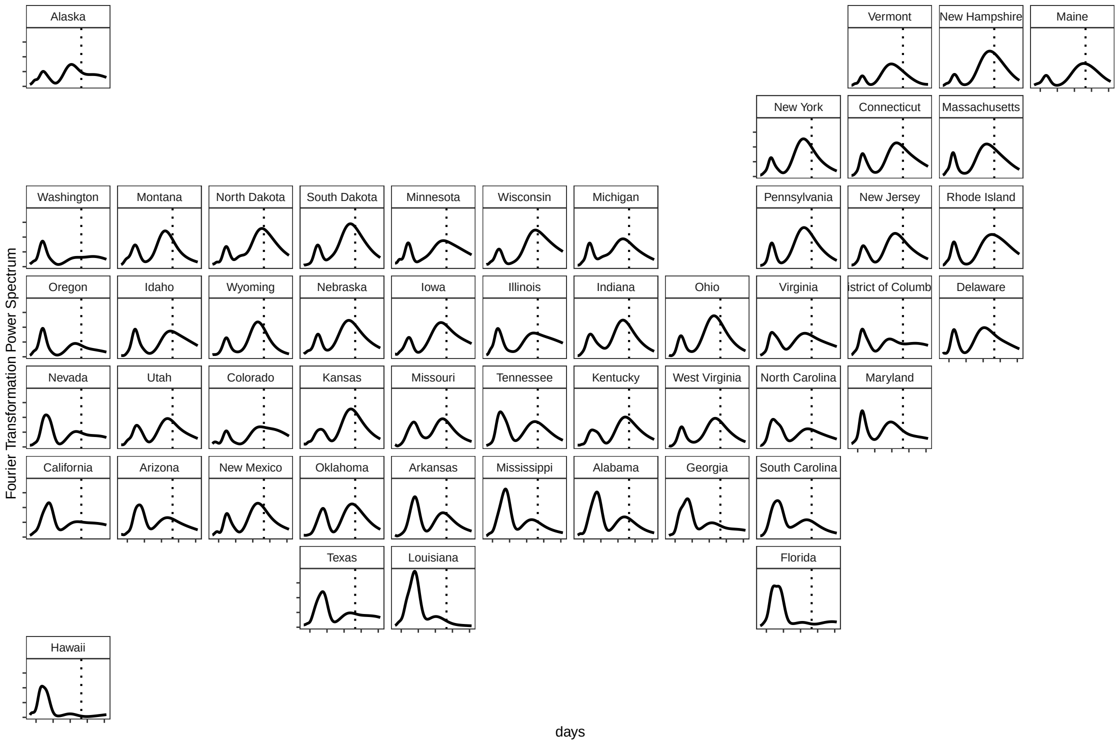


**Figure S2**: **United States map of global wavelet transformations.** Wavelet transformations were calculated for the log of COVID-19 incidence for each U.S. state. A dashed line is shown at 365 days for ease of identifying the power of the annual component of the global wavelet transformation.

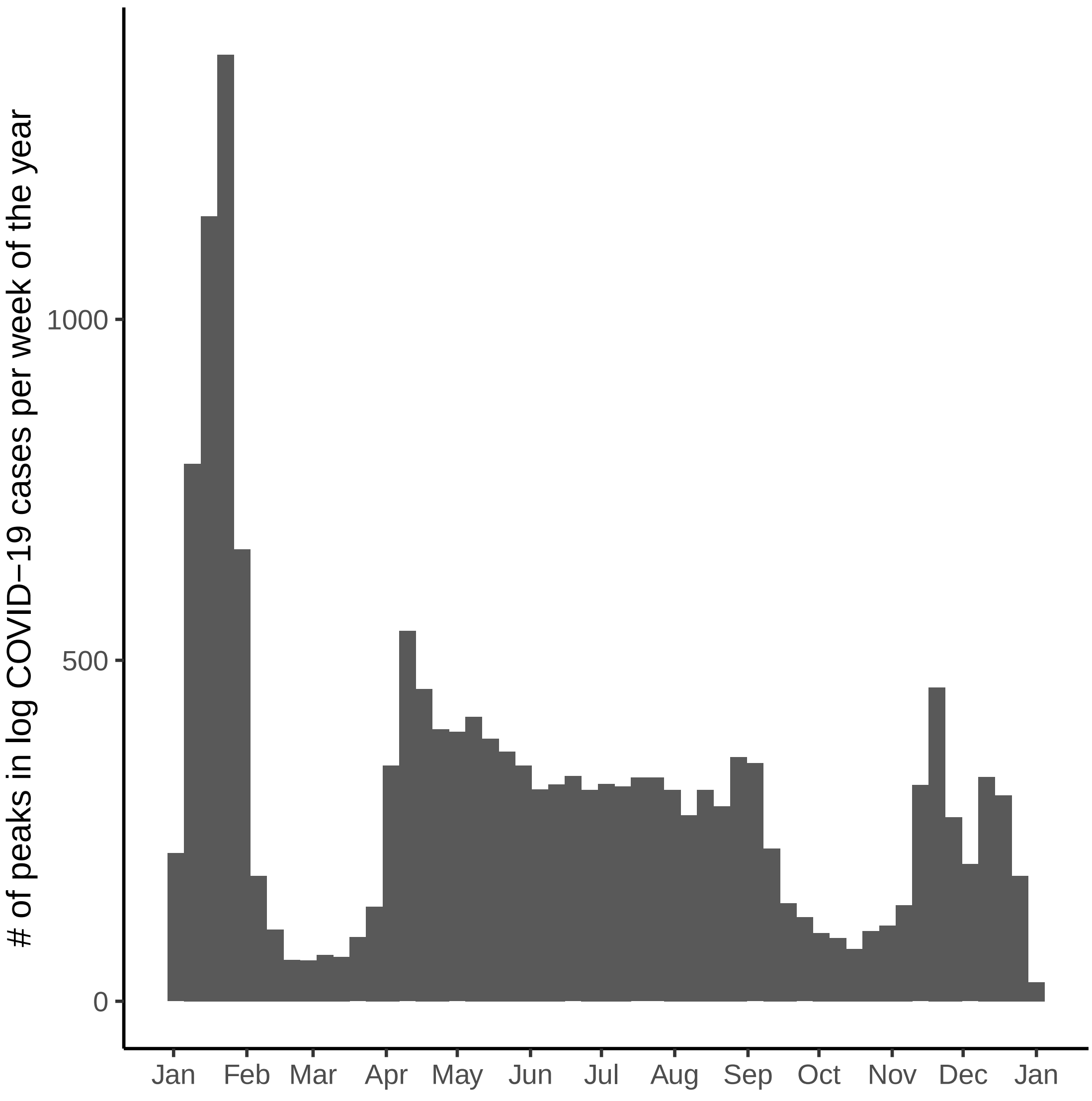


**Figure S3**: **Histogram of the timing of peaks in COVID-19 cases by week of the year.** Peaks in log COVID-19 cases were calculated for all FIPS in the United States as estimated by the NY Times (*12*) and are defined as any local maximum in the log of COVID-19 cases per 100k individuals with a minimum prominence of $10^3$. The histogram shows the number of peaks across all FIPS and the entire timeseries available that occurred during a given week of the year. This shows the regularity of the January winter wave, with less regularly timed waves occurring in the spring or summer and in the early winter.

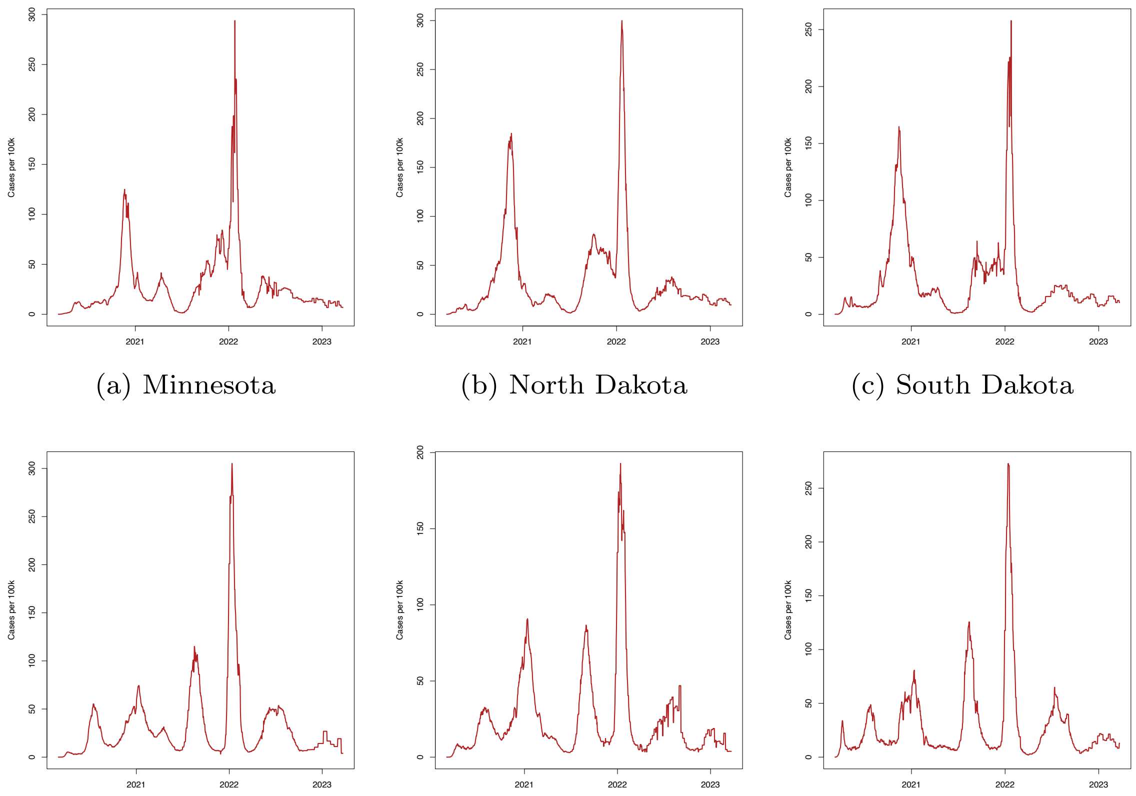


(a) Minnesota (b) North Dakota (c) South Dakota

(d) Florida (e) Georgia (f) Louisiana

**Figure S4**: **Cases of COVID-19 for the states with the coldest (a-d) and warmest (d-f) winter climates.** The number of cases of COVID-19 per 100k are shown for the same six states as shown in fig. S1

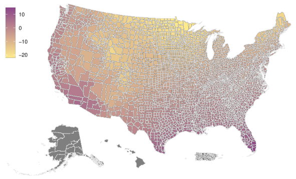

(a) Minimum temperature

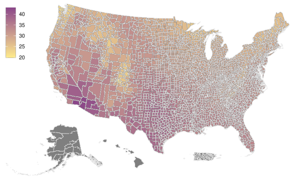

(b) Maximum temperature

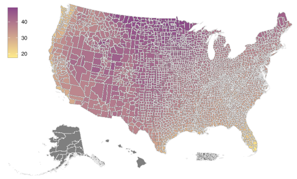

(c) Temperature difference

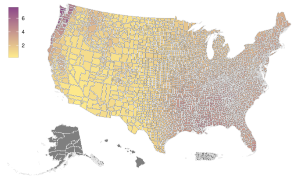

(d) Average precipitation

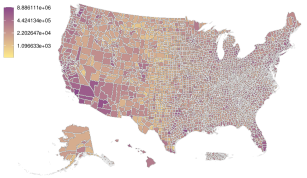

(e) Population size

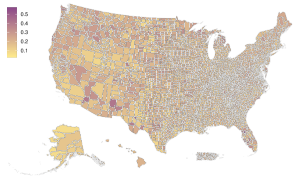

(f) Population 65+

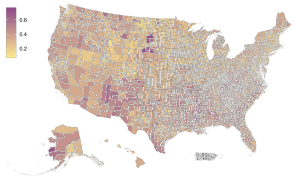

(g) Population in poverty

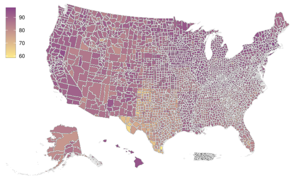

(h) Population insured

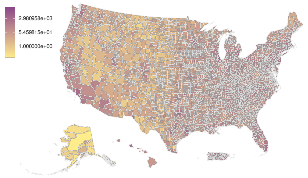

(i) Population density

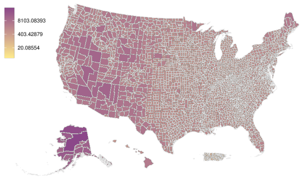

(j) Geographic area

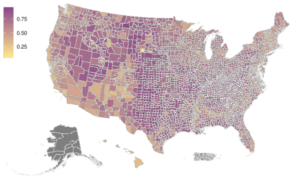

(k) 2020 Republican vote share

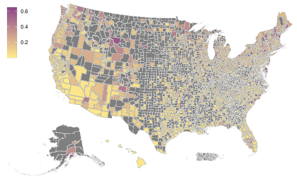

(l) Mobility (retail)

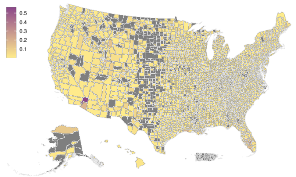

(m) Mobility (workplace)

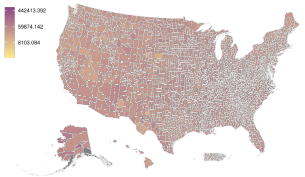

(n) Total cases COVID-19

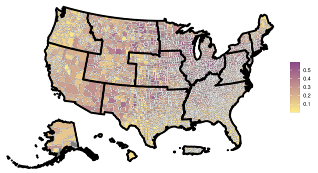

(o) COVID-19 annual periodicity with HHS regions

**Figure S5**: **County-level map of variables considered across the United States.** Maps of each variable used in comparison to the strength of the annual component of the COVID-19 epidemic at the county level. Where data is not available the county is shown in gray. Population size, population density, and county geographic area are colored on a log scale. For a detailed description of each variable and its source, please see the material and methods.

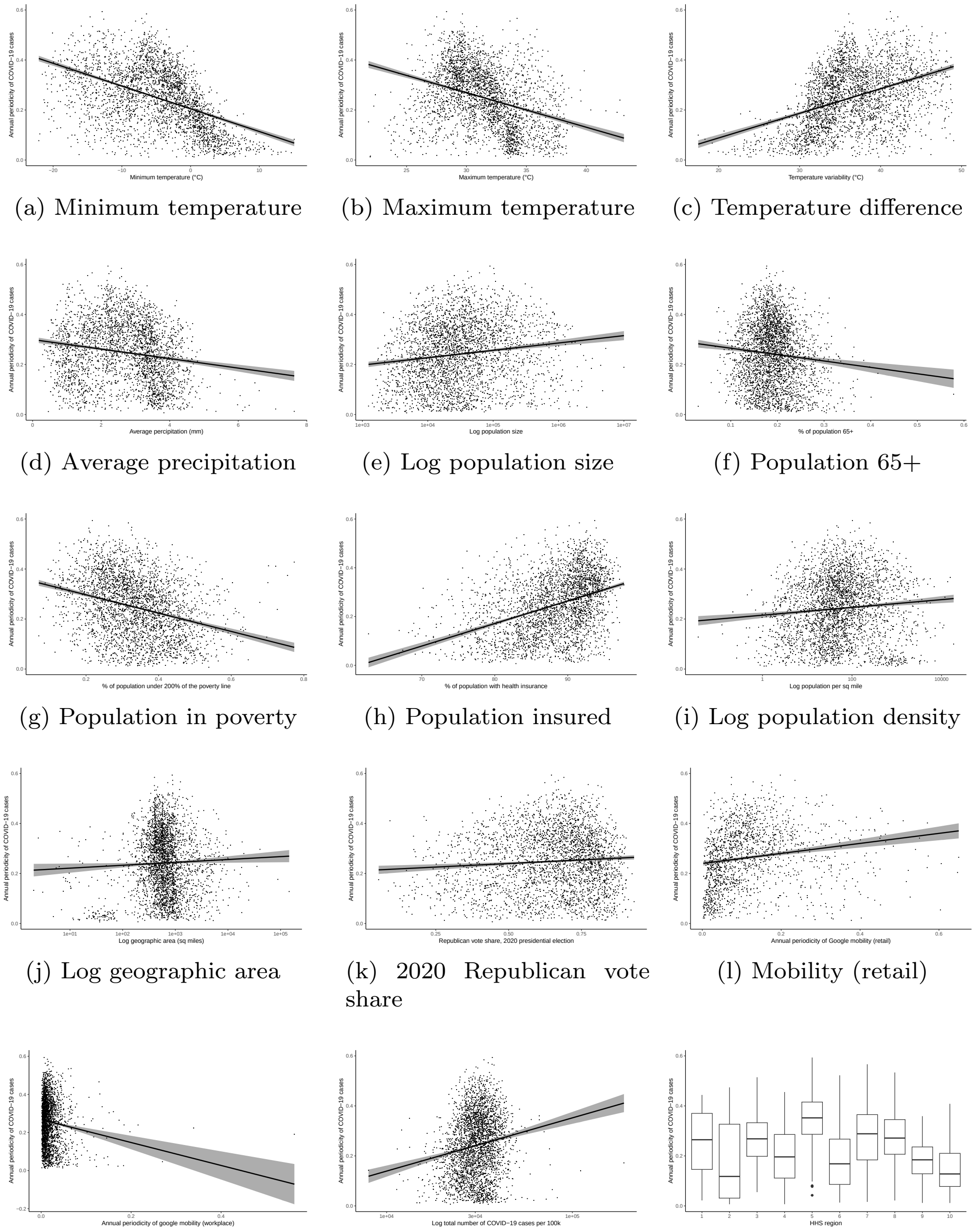

(a) Minimum temperature (b) Maximum temperature (c) Temperature difference

(d) Average precipitation (e) Log population size (f) Population 65+

(g) Population in poverty (h) Population insured (i) Log population density

(j) Log geographic area (k) 2020 Republican vote share (l) Mobility (retail)

(m) Mobility (workplace) (n) Log total COVID-19 cases (o) HHS region (ANOVA)

**Figure S6**: **Annual component of COVID-19 cases versus variables considered across the United States.** The annual component is defined as the value of the global wavelet transformation at a period of 365 days. Each point represents a county or county equivalent region across the United States and its territories. Lines represent a simple linear regression and it's 95% confidence interval. HHS regions are plotted using a box-and-whisker plot that show the 1st through 3rd quartiles. Because of the large sample sizes, all regressions are statistically significant. For a detailed description of each variable and its source, please see the material and methods.

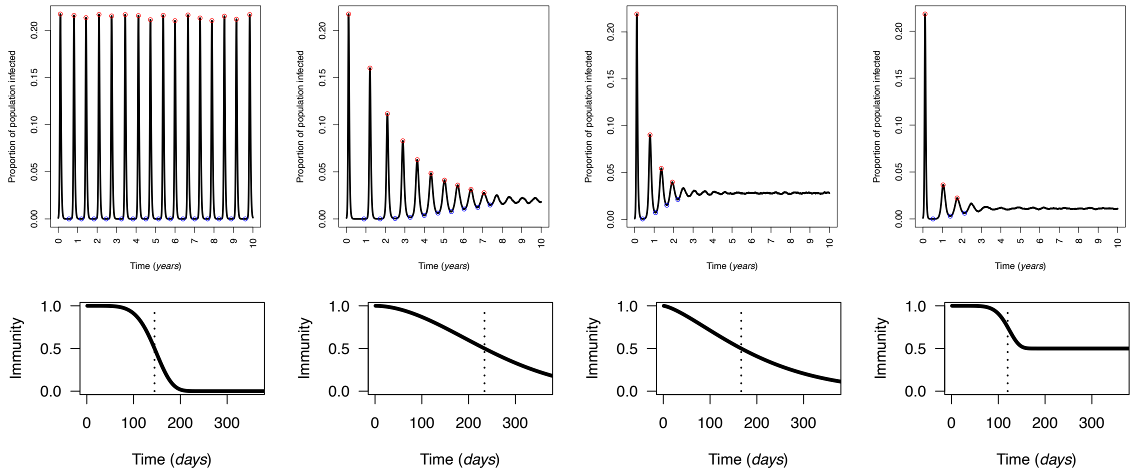


(a) Large $k$, small $\omega_\infty$

(b) Moderate $k$, small $\omega_\infty$

(c) Small $k$, small $\omega_\infty$

(d) Large $k$, large $\omega_\infty$

**Figure S7**: **Example simulated dynamics and waning immunity functions.** Four example individual-based simulations and the waning immunity functions that produced the dynamics. The top figure in each pair shows the number of newly infected individuals as a fraction of the total population. Red dots represent infection peaks and blue dots represent infection troughs as determined by our described algorithm. The bottom figure in each pair shows the individual waning immunity functions. Parameterization of each simulation are as follows: a) $\omega_\infty = 4.1 \cdot 10^{-4}$, $k = 5.4$, $\lambda = 144.8$; b) $\omega_\infty = 5.8 \cdot 10^{-4}$, $k = 1.9$, $\lambda = 234.0$; c) $\omega_\infty = 8.9 \cdot 10^{-6}$, $k = 1.4$, $\lambda = 166.6$; and d) $\omega_\infty = 0.5$, $k = 6.7$, $\lambda = 120.0$

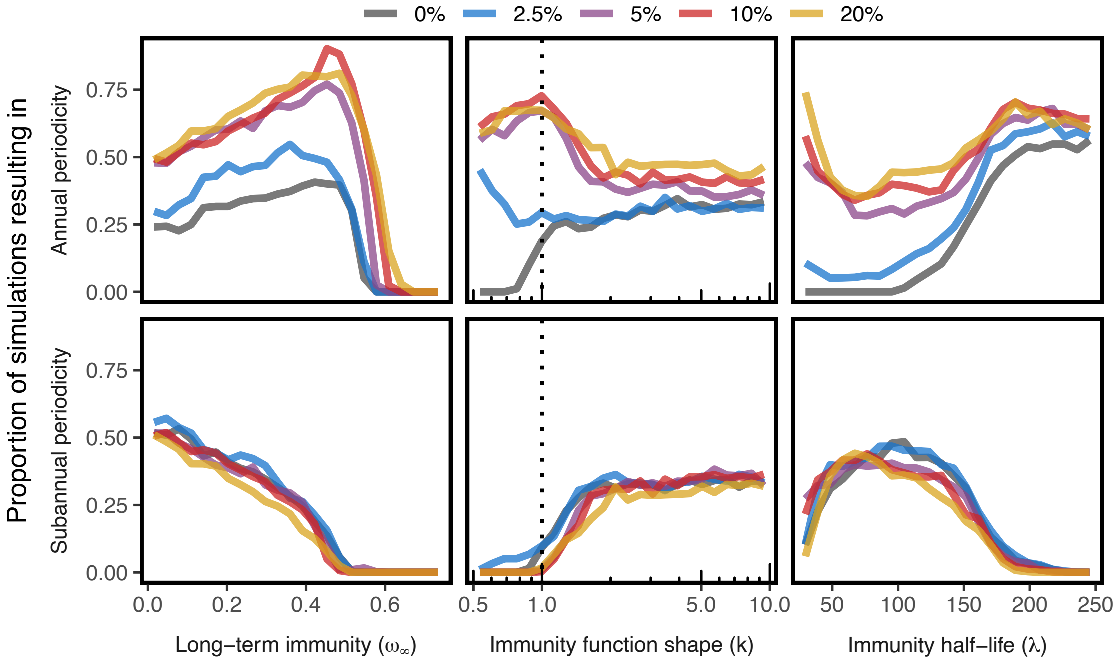


**Figure S8**: **Seasonal forcing simulations over only four years.** The same 125,000 simulations of the individual-based implementation of the SIRS model with individually waning immunity as described in the main text, but only the first four years of the simulation are analyzed. The 25,000 simulations for each of five different parameterizations of seasonal forcing are shown in color ($\theta$ = 2.5%, 5%, 10%, and 20%, or when the peak transmission rate is $\theta$% higher transmission than at its lowest). Immune parameters were chosen randomly with Latin Hypercube Sampling for each seasonality regime ($\omega_\infty \in (0, 0.75)$, $k \in (0.5, 10)$, and distributed logarithmically $\lambda \in (25, 250)$). Annual periodicity is defined as any simulation with an inter-epidemic period of between 9 and 18 months and sub-annual of less than 9 months, both requiring more than 3 total waves during the 4 simulation years to filter out strongly damped oscillations. Simulations that did not result in either annual or sub-annual waves either reached a stable equilibrium endemic state or resulted in waves less frequent than every 18 months. The dashed lines represent exponentially waning immunity, a good proxy for the classic SIRS model, and where undamped sub-annual waves are never generated. The results are not meaningly different than when the longer 10 year time period is considered as shown in Fig. 4.

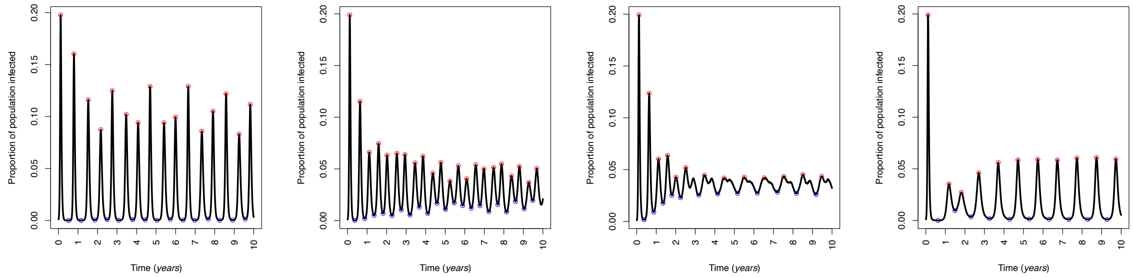


(a) Large $k$, moderate $\omega_\infty$ (b) Large $k$, moderate $\omega_\infty$ (c) Moderate $k$, small $\omega_\infty$ (d) Small $k$, moderate $\omega_\infty$

**Figure S9**: **Example simulated dynamics with seasonal forcing.** Four example individual-based simulations showing the complicated dynamics that can arise from the interaction of waning immunity and seasonal forcing. The top row shows two simulations that maintain sub-annual periodicity, though the magnitude of the waves is chaotic as a function of the seasonal forcing. The bottom row has two simulations in which the dynamics are "forced" onto an annual cycle. In C, the epidemic has sub-annual waves for the first few years, before settling into the annual cycle. This is perhaps an example of what could happen to the COVID-19 epidemic going forward Parameterization of each simulation are as follows: a) $\omega_\infty = 2.6 \cdot 10^{-1}$, $k = 4.3$, $\lambda = 137.2$; b) $\omega_\infty = 3.5 \cdot 10^{-1}$, $k = 8.3$, $\lambda = 91.4$; c) $\omega_\infty = 1.3 \cdot 10^{-1}$, $k = 1.8$, $\lambda = 107.2$; and d) $\omega_\infty = 2.2 \cdot 10^{-1}$, $k = 1.1$, $\lambda = 233.2$

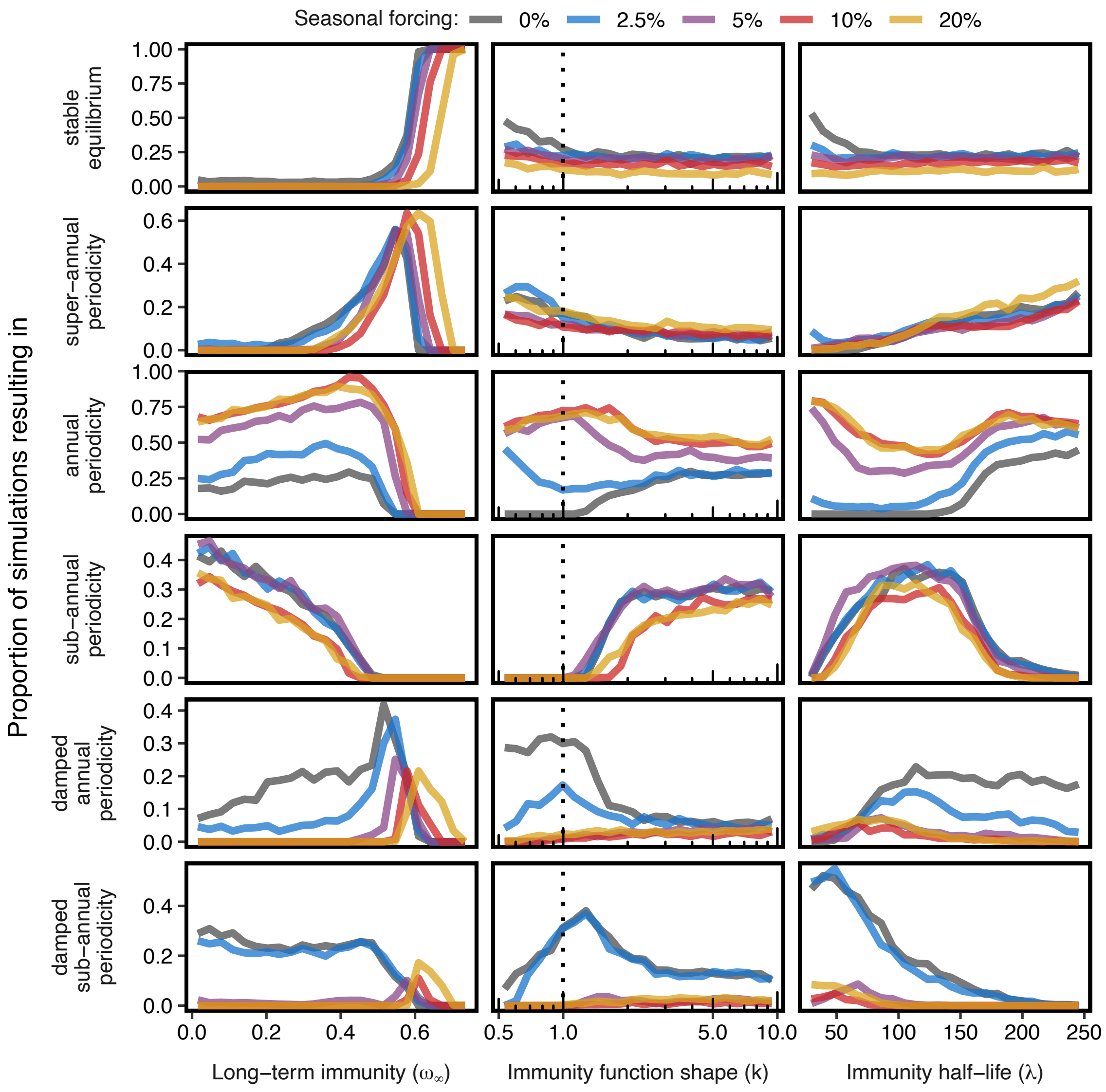


**Figure S10**: **Categorical periodic results of simulations.** The same 125,000 as in fig. 4 (10 simulation years) plotted to show other possible results than just annual and sub-annual periodicity. Annual periodicity is defined as any simulation with an inter-epidemic period of between 9 and 18 months and sub-annual of less than 9 months, both requiring more than 5 total waves during the 10 simulation years to filter out strongly damped oscillations. Those with 5 or fewer waves were considered to be damped. Super-annual periodicity is defined as simulations with an inter-epidemic period of greater than 18 months. Because of the possible long-term dynamics of these simulations, damped versus stable periodicity was not determined. Simulations that resulted in a single peak of infections are classified as stable equilibria. The dashed lines represent exponentially waning immunity, a good proxy for the classic SIRS model.

**Table S1**: **Multiple regression of county-level COVID-19 cases to all climatic and demographic variables.**

The results of a multivariable linear regression between COVID-19 cases and all numerical variables considered (except for temperature variability and population density as they are combinations of the minimum and maximum temperatures and population size and geographic area respectively). The table includes the coefficients for each variable in the model and their 95% confidence intervals. Variables with significant coefficients are bolded. The entire model is significant with the adjusted $r^2 = 0.541$

| Variable | Coefficient | 95% Confidence Interval |
|---|---|---|
| **Minimum temperature** | $\mathbf{-1.29 \cdot 10^{-2}}$ | $(-1.42 \cdot 10^{-2}, -1.15 \cdot 10^{-2})$ |
| **Maximum temperature** | $\mathbf{4.85 \cdot 10^{-3}}$ | $(2.17 \cdot 10^{-3}, 7.54 \cdot 10^{-3})$ |
| Average monthly precipitation | $-5.81 \cdot 10^{-3}$ | $(-1.18 \cdot 10^{-2}, 2.21 \cdot 10^{-4})$ |
| **Log population size** | $\mathbf{6.19 \cdot 10^{-2}}$ | $(4.94 \cdot 10^{-2}, 7.54 \cdot 10^{-2})$ |
| **% Population age 65+** | $\mathbf{0.208}$ | $(9.20 \cdot 10^{-2}, 0.324)$ |
| % **Population under 200% of the poverty line** | $\mathbf{-6.02 \cdot 10^{-2}}$ | $(-0.120, -2.95 \cdot 10^{-5})$ |
| **% Population insured** | $\mathbf{3.59 \cdot 10^{-3}}$ | $(2.37 \cdot 10^{-3}, 4.81 \cdot 10^{-3})$ |
| **Log geographic area** | $\mathbf{-6.54 \cdot 10^{-2}}$ | $(-7.78 \cdot 10^{-2}, -5.30 \cdot 10^{-2})$ |
| **Republican vote %, 2020 presidential election** | $\mathbf{0.248}$ | $(0.213, 0.284)$ |
| Google mobility periodicity (retail) | $-1.17 \cdot 10^{-2}$ | $(-6.51 \cdot 10^{-2}, 4.17 \cdot 10^{-2})$ |
| **Google mobility periodicity (workplace)** | $\mathbf{-0.268}$ | $(-0.440, -9.67 \cdot 10^{-2})$ |
| **Log total number of COVID-19 cases** | $\mathbf{0.320}$ | $(0.266, 0.373)$ |

**Table S2**: **Parameterization for simulations of SIR model with waning immunity.** The three parameters that govern the waning immunity function were varied based on Latin hypercube sampling. The ranges they were chosen from are given. The other parameters were fixed across all 125,000 simulations.

| Parameter | Value |
|---|---|
| $R_0$ | 2.5 |
| $1/\gamma$ | 7 days |
| $\omega_\infty$ | (0,0.75) |
| $k$ | (0.5,10) |
| $\lambda$ | (25,250) |
| Population size | 1e6 |
| Initial infected | 1 |
| Simulated time | 3650 days |
| $\theta$ | 0, 0.025, 0.05, 0.1, or 0.2 |
| $s_\theta$ | 0.5 |
